# Parental Perception of Children Sleep Pattern Changes During FIFA 2022

**DOI:** 10.64898/2025.12.21.25342765

**Authors:** Fadi Aljamaan, Abdullah Alhuzaimi, Shereen A. Dasuqi, Nasser Alharbi, Ibraheem Altamimi, Reem Alageel, Hadeel Alsulami, Amr Jamal, Shuliweeh Alenazi, Mohammed Alarabi, Khaled Saad, Elshazaly Saeed, Reema Alrabiah, Ahmad Alhadeed, Khalid Alhasan, Ahmed S. BaHammam, Mohamad-Hani Temsah

## Abstract

**Introduction:** The circadian clock is an internal, ∼24-hour biological timer that synchronizes physiology with the day-night cycle. Social jetlag (SJL) describes the misalignment between this internal clock and social schedules, a condition affecting approximately 70% of the population and linked to a spectrum of metabolic, mental, and cognitive health issues. This study examined how the 2022 FIFA World Cup disrupted normal children’s sleep routines and other associated factors from parents’ prospect.

**Methods:** An online, cross-sectional survey was distributed to parents (N=848). The questionnaire collected sociodemographic data, children’s habitual sleep habits, and changes perceived during the 2022 FIFA World Cup. SJL was defined as a ≥1-hour delay in bedtime on weekends versus weekdays. Multivariable logistic regression analyses identified factors associated with perceived sleep changes and SJL.

**Results:** Over half (53.4%) of the children exhibited pre-existing weekend SJL. Children aged 5-11 (OR=1.847, p<0.001), higher socioeconomic status (OR=1.383, p<0.001), international residency (OR=2.845, p<0.001) were significant predictors of baseline weekend SJL. During the tournament, 33% and 17.8% of parents reported their children had delayed sleep (≥1 hour) on weekdays and weekends due to match watching, respectively. Regression analysis revealed that these parental perceived FIFA impact on their children sleep was significantly associated with weekdays SJL OR= 1.958, p=0.001 and weekends OR= 1.784, p=0.009 during the FIFIA season.

**Discussion:** Our findings indicate that major social events can exacerbate circadian misalignment and SJL, likely driven by social conformation. The high prevalence of baseline SL confirms it is a widespread pediatric health issue. The vulnerability of the 5-11 age group shows SJL is not exclusively an adolescent problem.

**Conclusion:** Major sporting events significantly disrupt children’s sleep schedules, compounding the public health issue of chronic SJL. Proactive guidance for families is needed during such events. More importantly, these findings underscore the urgent need for structural changes to align social schedules with pediatric circadian biology.

## Introduction

The circadian clock is an internal, self-sustained ∼24-hour biological timer that synchronizes human’s physiology and behaviour with the day-night cycle, regulating fundamental processes from sleep and metabolism to hormone secretion[1]. This alignment allows human to anticipate daily recurring environmental changes. However, in modern society, a pervasive form of circadian disruption has emerged: social jetlag (SJL). SJL describes the misalignment between an individual’s innate biological time and their socially imposed schedule, quantified by the discrepancy in sleep timing between work/school days and free days[2–4]. It is estimated that 70% of the population experiences SJL of at least one hour, with almost half experiencing two hours or more, making it concern for public health[1].

The health consequences of this misalignment are extensive and particularly concerning for developing children and adolescents. Whereas the adverse effects of shift work and jetlag are widely recognized, SJL is a highly underestimated chronic stressor[1]. Physiologically, processes like glucose metabolism and blood pressure are under circadian control, and their disruption is linked to cardiometabolic risk. For instance, SJL has been significantly associated with unfavourable cholesterol levels and vascular health factors in children aged 8-10 years[3]. Furthermore, a recent cross-sectional study showed that children aged 2-8 with ≥1 hour of SJL had 66% higher odds of being overweight or obese[5].

Beyond metabolic health, the psychosocial impact is profound. SJL is positively associated with a range of mental health and behavioural issues. In preschool children, longer SJL (≥1 h/d) is linked to overall emotional and behavioural problems, emotional symptoms, hyperactivity, and conduct problems[6]. In adolescents, it is associated with irritable mood, anxiety, decreased academic and cognitive performance, and attention problems[2,7]. This is compounded by the biological “phase delay” of puberty, which pushes natural sleep times later, creating a direct conflict with early school start times[2,8]. This conflict is often exacerbated by the use of light-emitting electronic devices in the evening, the blue light from which interferes with circadian rhythms and is a reported correlate of stronger SJL[1,2].

Major international sporting events, such as the FIFA World Cup, represent a unique, large-scale social phenomenon that can act as a powerful, time-limited “social obligation.” These events force a restructuring of daily routines, with match schedules often extending into late evening hours, potentially displacing sleep. This study aimed to assess the parental perceptions of their children baseline social jetlag, sleep pattern changes and any additional social jetlag (SJL) during the FIFA World Cup and identifying key sociodemographic and factors associated with this social jetlag.

## Methodology

### Study design and setting

This study represents a secondary analysis of data originally collected for a previously published research examining sleep pattern changes among children during the 2022 FIFA World Cup[9]. In the original study, a cross-sectional, online survey was administered between 27 November-25 December 2022, coinciding with the FIFA World Cup Qatar 2022.

For the present manuscript, we focused specifically on parental perceptions of children’s baseline social jetlag, sleep disturbances during the tournament, and factors associated with additional sleep schedule shifts. The survey targeted parents residing in Saudi Arabia, sharing the same time zone as Qatar, as well as parents living in countries six hours ahead.

### Participants and recruitment

Participants were recruited using convenience sampling. Invitations linking to the survey were circulated through widely used social media platforms, including WhatsApp, X (previously Twitter), and Facebook. Eligible participants were parents or guardians residing in the specified regions during the study period. Before beginning the questionnaire, participants reviewed an electronic information sheet and provided digital informed consent. Responses were anonymous, and participation was voluntary.

Data collection was monitored daily and closed once the targeted sample was reached. A total of 848 parents completed the survey and were included in this study.

### Survey instrument and measures

#### Questionnaire structure

The survey captured sociodemographic information, children’s habitual sleep patterns, perceived changes during the FIFA tournament, and how families followed World Cup matches. The questionnaire also incorporated the validated Arabic version of the Children’s Sleep Habits Questionnaire (CSHQ) to assess baseline sleep behaviours.

Parents reported weekday and weekend bedtimes under usual conditions and during the FIFA period. Additional questions explored perceived changes in sleep duration, general sleep behaviour during the tournament, and viewing preferences (live vs. delayed match viewing or not watching).

### Definition of social jetlag

SJL was defined based on discrepancies in sleep timing:

- Baseline SJL: ≥1-hour delay in weekend bedtime compared with weekday bedtime.
- FIFA-related weekday SJL: ≥1-hour delay in weekday bedtime during the World Cup compared with usual weekdays.
- FIFA-related weekend SJL: ≥1-hour delay in weekend bedtime during the tournament compared with usual weekends.
- All SJL outcomes were dichotomized (yes/no).

### Socioeconomic status index (SESi)

A composite SES index was calculated through categorical factor analysis incorporating parental education, employment, household size, and perceived socioeconomic standing. Indicators with factor loadings ≥0.32 on the latent factor were retained, and the resulting SES index was standardized.

### Ethical Approval Statement

The research study was reviewed and approved by the Institutional Review Board at King Saud University, Riyadh, Saudi Arabia (reference# 19/0953/IRB).

### Statistical data analysis

The mean and standard deviations were used to describe continuous variables which showed statistical normality and the medians and inter-quartile ranges were used to describe continuous variables that showed statistical normality assumption violations. The statistical normality assumption for the metric variables was assessed with the histograms and the statistical normality Kolmogrove-Smirnove (KS) tests.

The frequency and percentages were used to describe the categorically measured factors. The Variance Inflation Index (VIF) and Tolerance collinearity diagnostic tests were used to assess collinearity between analysed variables. The multiple response dichotomies analysis was used to describe the variables measured with more than options. The categorical Factor Analysis was used to reduce the parental socioeconomic and educational factors into a standardized socioeconomic factor index (SESi) via regressing the parents (educational level, households’ size, employment and gent socioeconomic factor state score, a salient item-factor loading in the factor analysis was defined as loading of each indicator to the latent SESi factor of ≥ 0.32 points. The children’s SJL was computed via estimating the time difference between weekend and weekdays sleep times, a child with delayed sleep time ≥1 hours during weekends vs weekdays were considered to have been positive for (SJL), also sleep jetlag related (SJL) due to football games days was compared against weekends and weekdays sleep times in the same manner. Multivariable Logistic Binary Regression analysis was applied to dichotomous measured sleep related outcomes and the association between the predictor independent variables in the multivariate Logistic Binary regression analysis was expressed as an Odds Ratio (OR) with their associated 95% confidence intervals. The SPSS IBM statistical computing program version 21 was used for the statistical data analysis, and alpha significance level was considered at 0.050 level.

## Results

Eight hundred and forty-eight parents participated in the study and completed fully the online questionnaire. Table 1 displays the descriptive analysis for parents and household sociodemographic characteristics. Mothers comprised 60.6% of the participants. Parental age distribution was as follows:15.3% aged 20-34 years, 44.5% 35-44 years, 27.9% 45-54 years and 12.3% aged >=55 years. The educational level for the parents, 81.1% of the parents had a university degree or higher educational levels, while the rest achieved school education only.

**Table. 1:** Descriptive analysis of the parents’ sociodemographic characteristics. N=848.

| Variable | Frequency | Percentage |
| --- | --- | --- |
| **Sex** |  |  |
| Female | 514 | 60.6 |
| Male | 334 | 39.4 |
| **Age group** |  |  |
| 20-34 | 130 | 15.3 |
| 35-44 | 377 | 44.5 |
| 45-54 | 237 | 27.9 |
| 55-64 or older | 104 | 12.3 |
| **Educational Level** |  |  |
| Primary school | 13 | 1.5 |
| Middle school | 24 | 2.8 |
| Highschool | 123 | 14.5 |
| University or postgraduate degree | 688 | 81.1 |
| **Employment** |  |  |
| Unemployed/Housewife/students | 247 | 29.1 |
| Freelance job | 62 | 7.3 |
| Employed | 539 | 63.6 |
| **Number of children in the family (household's size), median (IQR)** |  | 3 (3) |
| **Do you have a child whose age is 5-11 years?** |  |  |
| No | 247 | 29.1 |
| Yes | 601 | 70.9 |
| **Do you have a child whose age is 12-18 years?** |  |  |
| No | 329 | 38.8 |
| Yes | 519 | 61.2 |
| **Has this questionnaire encouraged you to observe symptoms of poor sleep in yourself or your family?** |  |  |
| No | 291 | 34.3 |
| Yes | 557 | 65.7 |
| **Place of residence** |  |  |
| Saudi Arabia | 751 | 88.6 |
| Other countries=International | 97 | 11.4 |

Regarding the parental employment status, 29.1% were unemployed (housewives or students), 7.3% had a freelance job and 63.6% were employed. The median number of children in the family was 3 with an inter-quartile range of 3 children as well. The children age range was as follows: 70.9% of the parents had at least one child aged 5-11 years, and 61.2% of them had at least a child aged between 12-18 years.

Most participating parents were from Saudi Arabia 88.6%, while the rest were from other international participating sites whose time Zone was +6 hours from the Saudi or Qatari time Zone (Saudi Arabia and Qatar share same Time Zone).

After assessing the children’s usual bedtime during weekdays and weekends the (SJL) was defined as having difference in bedtime between weekends vs weekdays of >=1 hour. Most of the parents 53.4% admitted that their children had (SJL) in weekends routinely compared to weekdays. While 33% of the parents perceived that their children had (SJL) in weekdays compared to their norm during FIFA games, and 17.8% of them perceived (SJL) during FIFA weekends compared to their norm. The total usual slept hours by the children according to their parents was mean = 9.10 hours, SD=1.78 hours.

64.2% of the participating parents did not perceive that their children’s sleep hours had changed during the FIFA soccer matches period, while 25.5% of noticed a little change, 10.4% perceived a substantial change.

Moreover, 66.3% of the parents believed that FIFA had no impact on their children’s sleep in general, while 25.5% believed it had little and 5.2% believed it had mild impact but 2.2% and 0.8% respectively believed the FIFA season had moderate and high impact.

Figure-1 shows the breakdown of the local and international parental perceived FIFA impact on their children’s sleep, international parents had more high and moderate perception of the impact compared to local parents residing in KSA.

**Figure 1:**
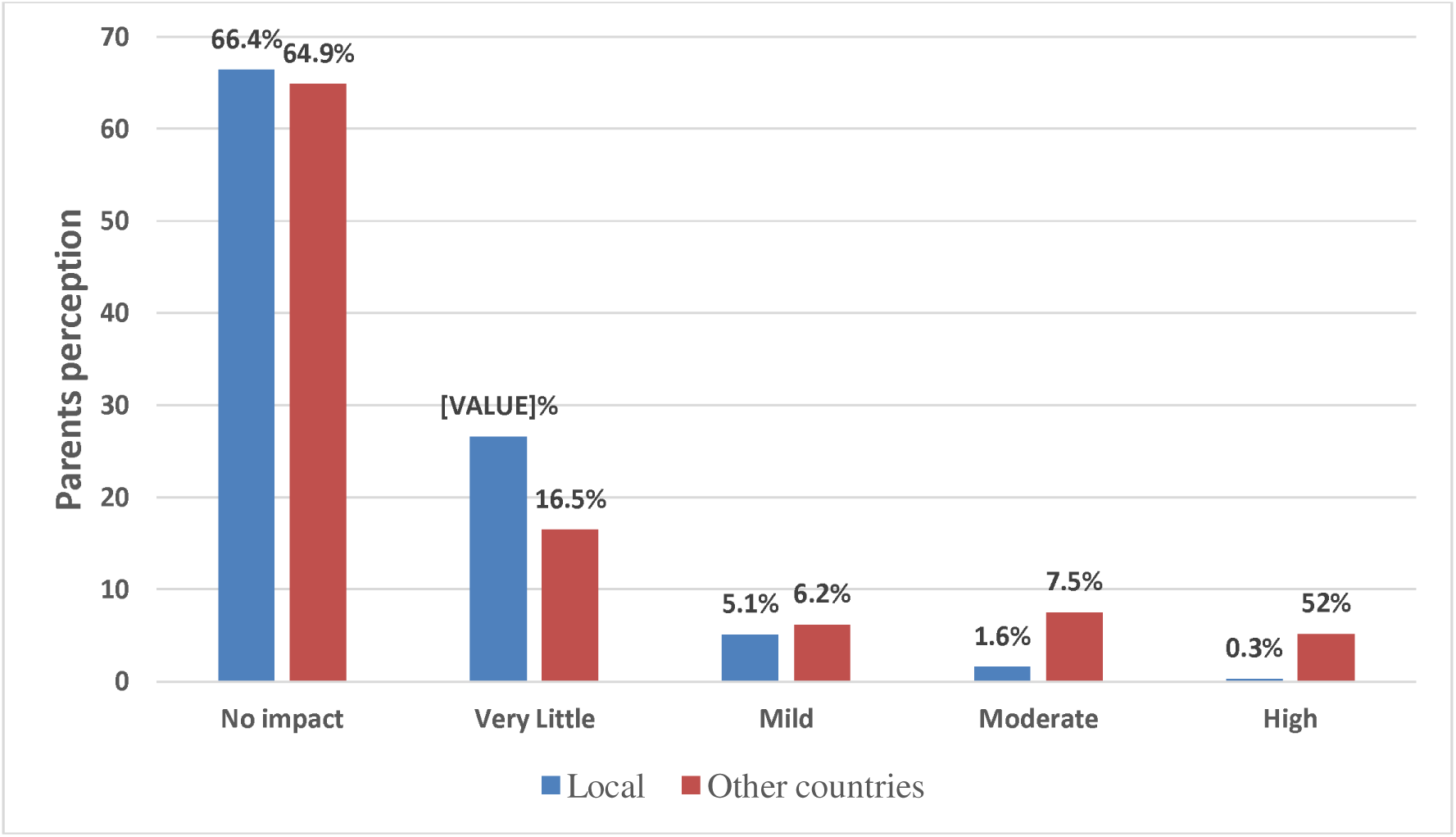
Parents’ perception of FIFA on their children’s overall sleep pattern.

**Table 2:** Parents perception of their children’s sleep habit change during FIFA matches.

| Social Jetlag during weekends vs weekdays |  |  |
| --- | --- | --- |
| No | 395 | 46.6 |
| Yes | 453 | 53.4 |
| Social jetlag during Football matches weekdays |  |  |
| No | 568 | 67 |
| Yes | 280 | 33 |
| Social jetlag during Football matches weekends |  |  |
| No | 697 | 82.2 |
| Yes | 151 | 17.8 |
| Total usual daily slept hours per nights, mean (SD) |  | 9.10 (1.78) |
| Did your child's sleep hours change due to watching the World Cup matches? |  |  |
| Never | 544 | 64.2 |
| A little | 216 | 25.5 |
| A lot | 88 | 10.4 |

The majority preferred live watch of the games either on TV or mobile phones, parents and children (42.7%,38.1%) respectively, while 26.7% of the parents admitted that they do not watch FIFA games and 17.6% for children. On the other hand, minority of both preferred watching FIFA football games offline 8.5% for parents and 5.7% for children as shown in Figure 2.

**Figure 2:**
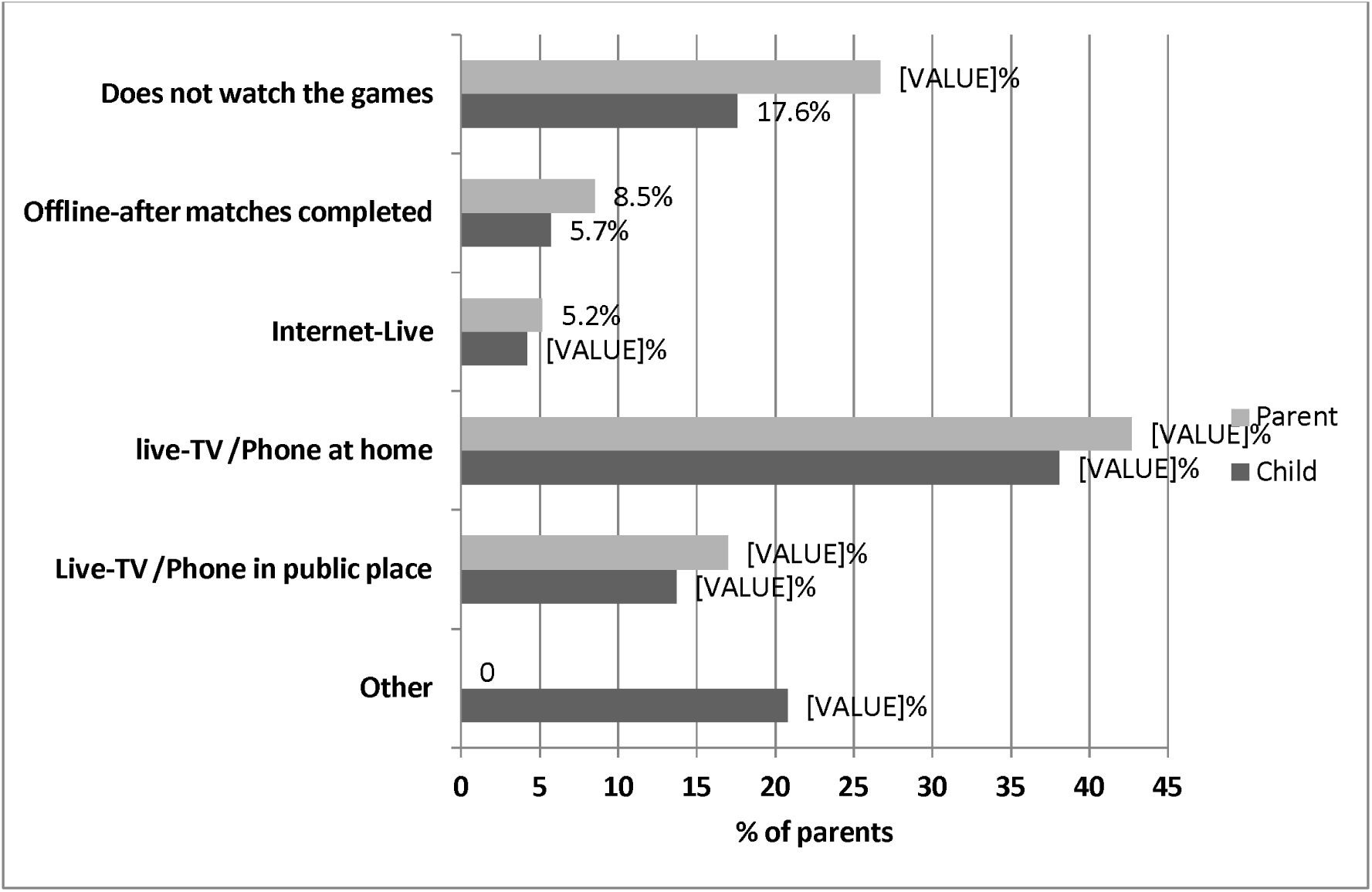
The parent and child used methods of watching FIFA matches.

Table 3 sheds light on multivariable Logistic Binary Regression Analysis of perceived parental variables significantly contributing to their children’s (SJL) during weekends compared to weekdays in general. Households socioeconomic state index (SESi) correlated significantly and positively with weekend (SJL) (1.38 times more for each 1 point of index, p-value<0.001). International parents compared to Saudi parents had significantly higher perception of weekend (SJL) for their children (2.845 times more, p-value<0.001). Parents of children aged between 5-11 years had higher perception of their children to have weekend (SJL) compared to other parents (1.847 times more, p-value<0.001). Parents who reported a change in their children’s sleep during FIFA season had higher perception of (SJL) during weekends in general regardless of FIFA (1.418 times more, p-value=0.003), while parental rated FIFA impact on their children sleep did not correlate with their perceived weekend (SJL) in general p-value=0.717.

**Table 3:** Multivariate Logistic Binary Regression Analysis of parental perception of their children’s (SJL) during weekends*.

|  | Multivariate Adjusted Odds Ratio | OR 95% C.I. |  | *p-value* |
| --- | --- | --- | --- | --- |
|  |  | Lower | Upper |  |
| **Sex of parent**<br>**= Male** | .845 | .590 | 1.209 | .356 |
| **Parental age** | .873 | .738 | 1.033 | .114 |
| **Households socioeconomic status (SESi)** | 1.383 | 1.162 | 1.646 | <0.001 |
| **Location**<br>**= International** | 2.845 | 1.719 | 4.709 | <0.001 |
| **Family with children aged 5-11 years** | 1.847 | 1.346 | 2.536 | <0.001 |
| **Parental perceived FIFA impact on their children's sleep**<br>**Yes** | 1.418 | 1.129 | 1.781 | .003 |
| **Weekend (SJL) during FIFA season**<br>**Yes** | .408 | .276 | .602 | <0.001 |
| **Constant** | .057 |  |  | <0.001 |

***Dependent outcome variable: parental perception of their children's (SJL) during weekends compared to weekdays in general No/Yes***
***\*Generally, apart from FIFA season***

Interestingly, parents who reported weekend (SJL) for their children during FIFA season were significantly less predicted to report weekend (SJL) in general regardless of the FIFA (59.2% times less, p-value<0.001).

Table 4 shows the multivariable Binary logistic regression analysis of the variables associated with parental perception of their children’s sleep change during FIFA season Mondial. Parents aged>=45 years had perceived significantly higher sleep change among their children during FIFIA season (1.307 times higher, p-value=0.004). Children’s baseline weekend (SJL) did not correlate significantly with their odds of sleep pattern change during FIFIA season, but children who had weekdays (SJL) during FIFIA season were perceived significantly by their parents to have sleep pattern change during the Mondial (1.958 times more, p-value=0.001). Likewise, the children who experienced (SJL) during FIFA weekend were perceived by their parents more significantly to have sleep pattern change during the Mondial (1.784 times more, p-value=0.008).

**Table 4:** Multivariate Logistic Binary Regression Analysis of parental perception of their children’s sleep pattern change during FIFA season.

|  | Multivariate Adjusted Odds Ratio | OR 95% C.I. |  | *p-value* |
| --- | --- | --- | --- | --- |
|  |  | Lower | Upper |  |
| **Sex of parent =Male** | .739 | .499 | 1.095 | .132 |
| **Parental age >=45** | 1.307 | 1.089 | 1.568 | .004 |
| **Location = International** | 1.324 | .793 | 2.209 | .283 |
| **Households socioeconomic status ( SESi)** | .943 | .784 | 1.135 | .536 |
| **Social jetlag\*** | 1.147 | .779 | 1.689 | .486 |
| **(SJL) in weekdays during FIFA season**<br>**Yes** | 1.958 | 1.319 | 2.906 | .001 |
| **(SJL) in weekends during FIFA season**<br>**Yes** | 1.784 | 1.158 | 2.748 | .009 |
| **Children watching football matches online after games already finished** | 0.271 | 0.124 | 0.593 | .001 |
| **Children who do not prefer watching Football** | 0.155 | 0.084 | 0.286 | <0.001 |
| **Period of the FIFA games** | 1.132 | 1.035 | 1.238 | .006 |
| **Constant** | 0.749 |  |  | .647 |

***Dependent outcome variable parental perception of their children's sleep pattern changes during FIFA season No/Yes***
**\*Parental perceived (SJL) of their children during weekend compared to weekdays apart of FIFA season**

On the other hand, children who watched the FIFA games later when the games had finished perceived by their parents significantly to have less effect on their sleep pattern during the Mondial (72.9% times less, p-value=0.001), similarly, children who do not prefer football match watching had significantly less effect on their sleep pattern during the Mondial (84.5% times less, p-value<0.001). The parental perception of their children’s sleep pattern changes during the Mondial had significantly increased along the Mondial progress period of FIFA games (OR 1.132, P value .006) Figure 3, shows that as the FIFA season had lapsed the parental mean predicted probability of being concerned about their child sleep pattern change tended to rise incrementally and steadily until the last few days of the Mondial.

**Figure 3.**
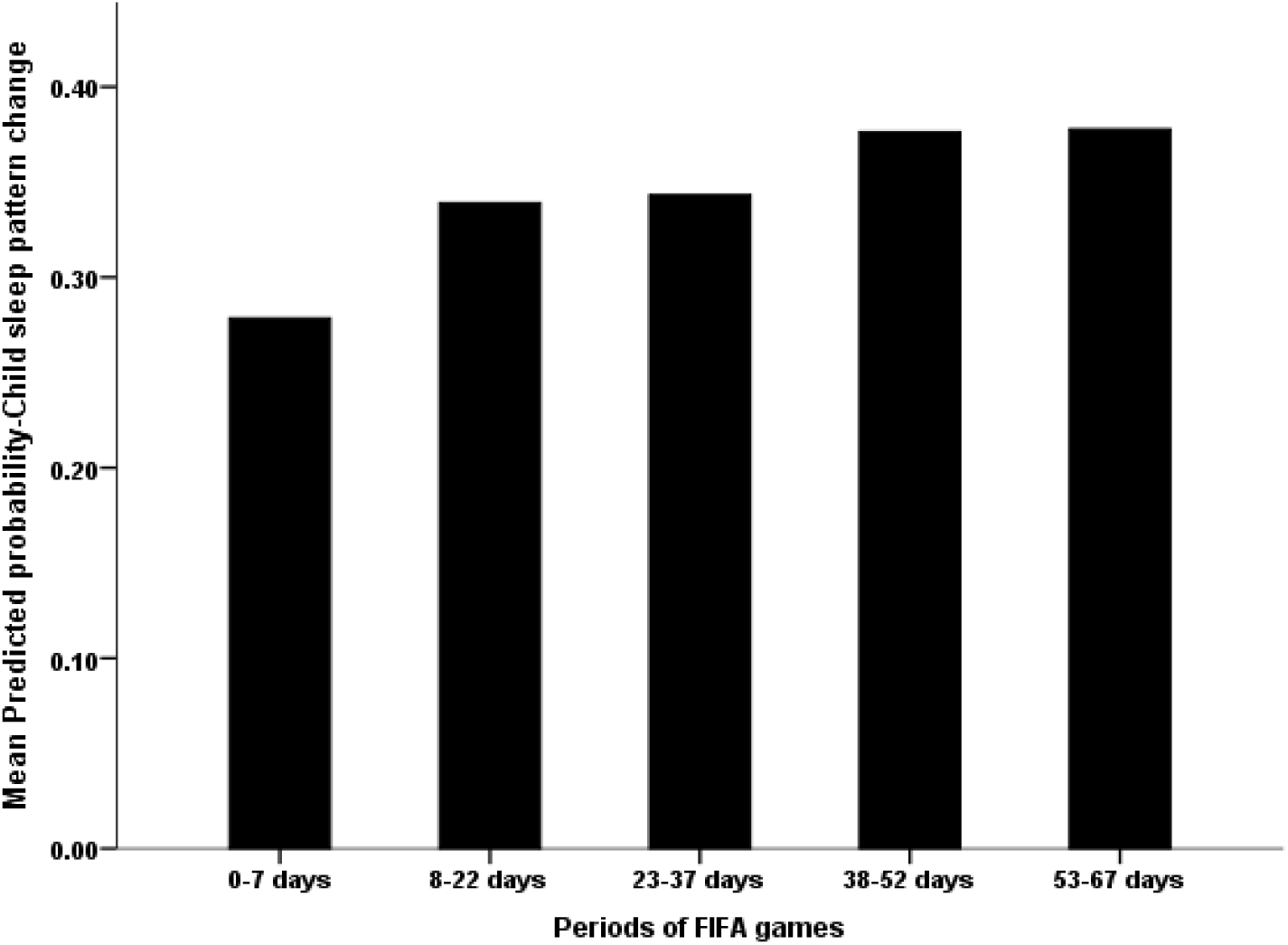
Mean parental perceived sleep pattern change during FIFA match during the Mondial season.

Table 4 sheds light on the variables associated with parental perceived (SJL) in FIFIA weekends. Parents aged >=45 years were significantly more predicted perceived weekends (SJL) during FIFA season (1.659 times more, p-value=0.033). While parents who had children aged 5-11 years perceived significantly less to have weekends (SJL) during FIFA (OR 0.625, p-value=0.022). Children whom their parents perceived their sleep pattern/habit changed during FIFA season had significantly more weekends (SJL) during FIFA season (1.403 times more, p-value=0.0130. The FIFA season progress or matches timing did not correlate with children’s weekends (SJL) during the Mondial.

**Table 4:** Multivariate Logistic Binary Regression Analysis for parental perception of their children’s (SJL) in weekend during FIFA season.

|  | Multivariate Adjusted Odds Ratio | OR 95% C.I. |  | p-value |
| --- | --- | --- | --- | --- |
|  |  | Lower | Upper |  |
| **Sex of parent**<br>**= Male** | .909 | .734 | 1.125 | .380 |
| **Parental age**<br>**$\geq 45$** | 1.659 | 1.043 | 2.638 | .033 |
| **Households' socioeconomic status (SESi)** | 1.025 | .819 | 1.284 | .828 |
| **Location**<br>**= International** | .645 | .337 | 1.233 | .185 |
| **Having children aged 5-11 years** | .625 | .419 | .934 | .022 |
| **Parental children perceived change in sleep pattern during FIFA season** | 1.403 | 1.074 | 1.833 | .013 |
| **Constant** | .283 |  |  | .192 |

***Dependent outcome variable= Parental perception of their children's (SJL) in weekend during FIFA season No/Yes.***

The variables associated with children’s weekdays SJL during FIFA season are shown in Table 5 (Multivariable Binary Logistic Regression). International parents had perceived significantly higher children’s weekdays (SJL) delay during FIFA (2.073 times higher, p-value=0.002). Parents having children aged 5-11 years had perceived significantly their children to have weekdays (SJL) during FIFA season (1.422 times more, p-value=0.047). Parents who reported that their children’s sleep pattern or behaviour changed during the Mondial had perceived that their children had significant (SJL) during FIFIA weekdays (1.758 times more, p-value=0.003). Children who had weekend (SJL) during FIFA season had significant incidence of (SJL) during weekdays also (4.004 times more, p-value<0.001).

**Table 5:**
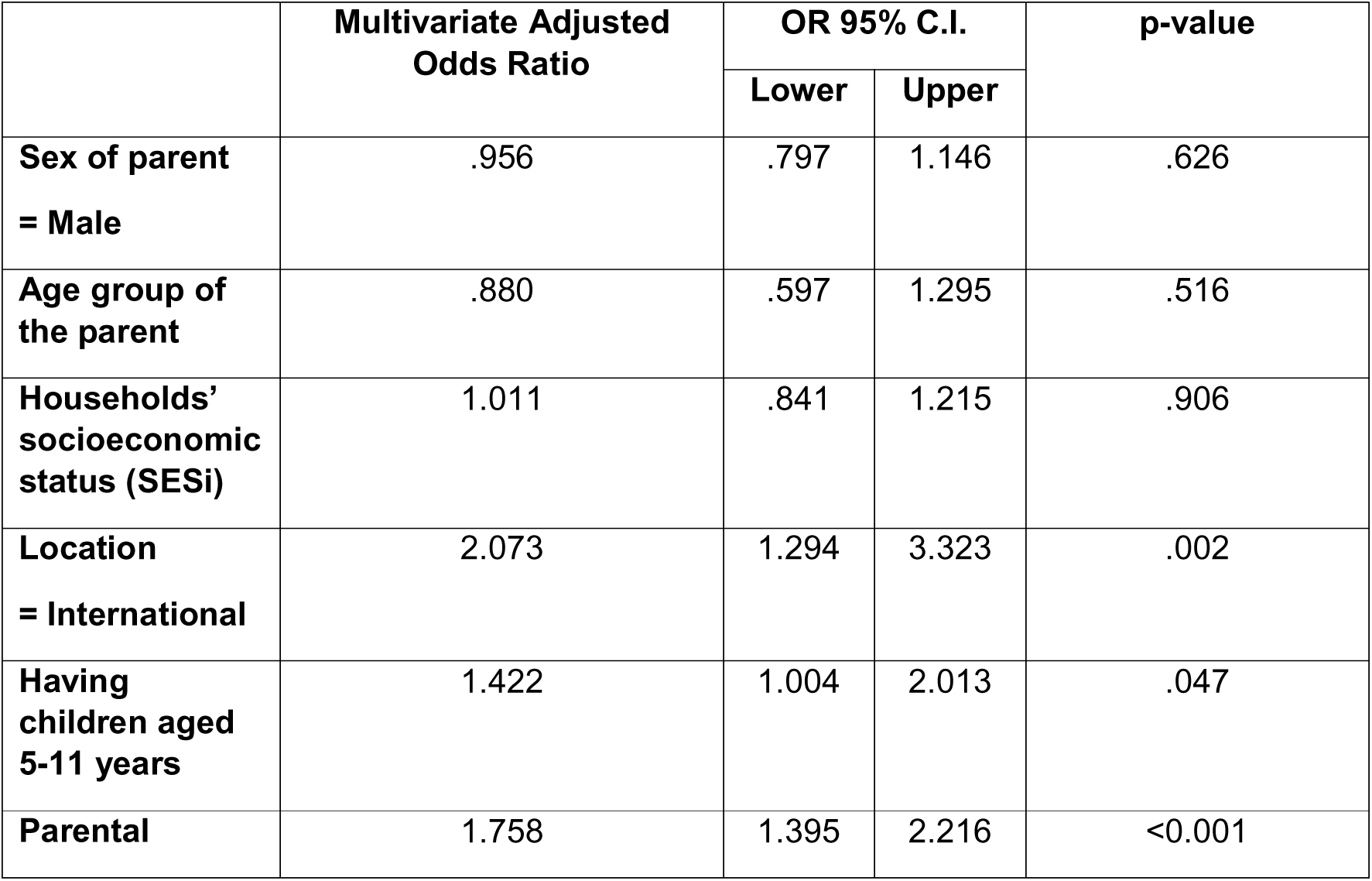
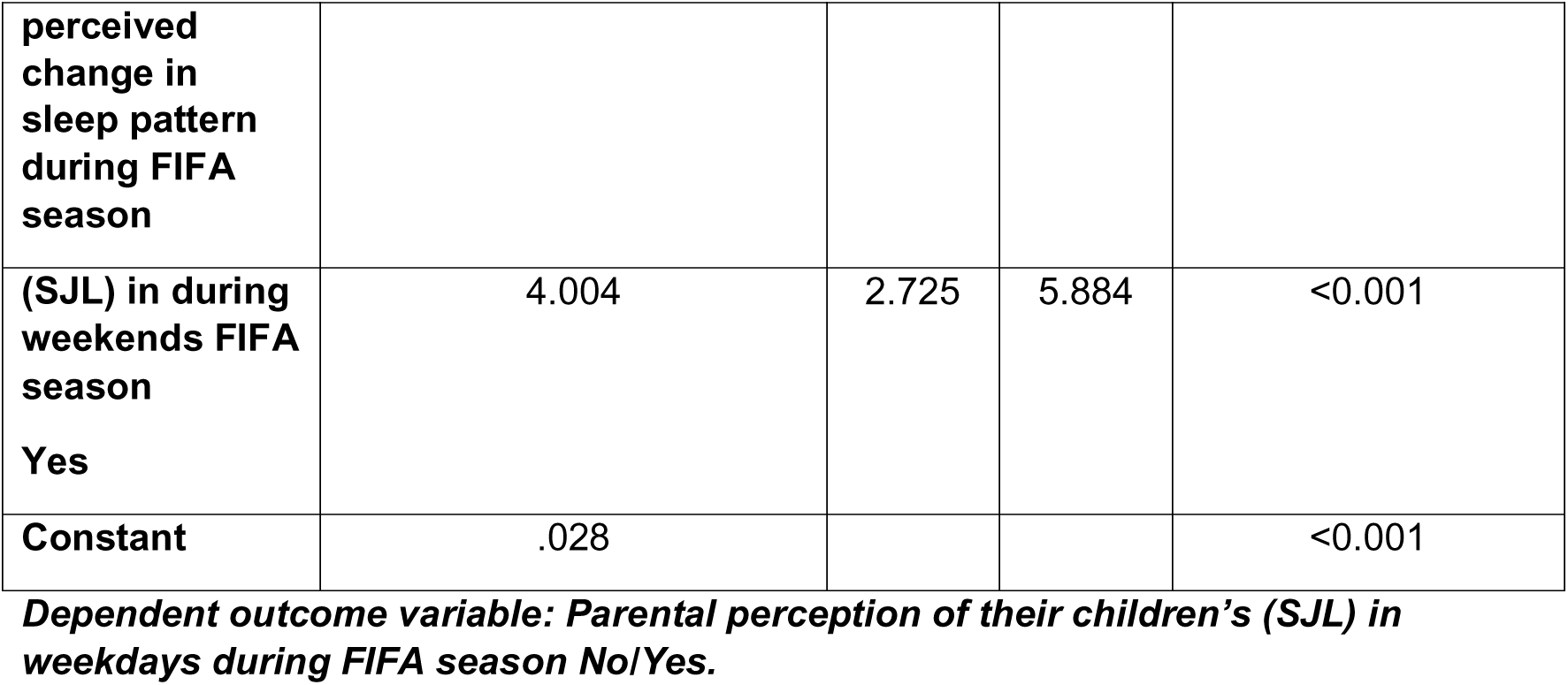
Multivariate Logistic Binary Regression Analysis of parental perception of their children’s (SJL) in weekdays during FIFA season.

## Discussion

This study provides a multifaceted examination of how the FIFA World Cup, a major globally synchronized social event, interacts with and exacerbates the universal public health issue of sleep disturbance in children. The results paint a concerning picture: a high baseline rate of chronic circadian misalignment within the pediatric population, which is then acutely and significantly worsened by the social obligation of late-night football match viewing. This obligation can be understood as a form of social pressure, which motivates conformity through a “sense of should”; a felt need to meet social expectations to stabilize interactions and reduce the cognitive costs of uncertainty, operating independently of explicit rewards or punishment[10].

Our finding of 53.4% parents observing that their children exhibited a baseline weekend SJL is a clear confirmation that circadian misalignment is not a rare or adult-centric issue, but a common condition of modern childhood. This high baseline prevalence of SJL observed in our sample indicates that a significant portion of children are already on this risky path, even before the added disruption of a major sporting event. This challenges the common belief that SJL is exclusively a problem of teenagers. It indicates that the displacement of sleep-wake times starts early, often influenced by parental habits and family routines[11].

The SJL phenomenon is exacerbated by what has been termed “weekend oversleeping” or “catch-up sleep,” a common behaviour where individuals extend sleep on free days to compensate for debt accumulated during the week[12]. While weekend catch-up sleep has been associated with some better health outcomes in adults[13]. This pattern maintains SJL by creating a weekly cycle of sleep deprivation and late sleeping, making it physiologically difficult to re-adjust to an earlier bedtime on Saturday night. This “Sunday morning jetlag” is a weekly reality for a lot of children, leading to excessive daytime sleepiness, which itself is chronotype-dependent[12]. This aligns precisely with literature, stating that most of the population experiences this form of misalignment, with almost half experiencing two hours or more social jetlag. The normalization of this chronic state is alarming which is manifested in our study by the finding that parents who reported SJL during FIFIA weekends tended to underscore social jetlag in general and consider this normal phenomenon. On the other hand, parents who perceived FIFA’s impact on their children sleep pattern being serious were also concerned and reported SJL in usual days apart from FIFA season.

SJL represents a persistent stressor on physiological systems that are under tight circadian control[14]. The wide-ranging health consequences identified in the literature underscore the seriousness of our findings. In the short-term, SJL is linked to poor and shortened sleep, impaired alertness, poor performance, mental health, hypertension, and an abnormal inflammatory status[15,16]. The long-term prospects are even more concerning, as SJL is associated with a higher risk of developing obesity, diabetes mellitus, metabolic syndrome, cancer, cardiovascular diseases, and cognitive impairments[1]. The mechanisms underlying associations between SJL and metabolic disorders are rooted in fundamental physiology. Processes such as glucose metabolism and blood pressure exhibit robust circadian rhythms. When behavioural cycles (sleep/wake, fasting/eating) are desynchronized from this internal clock, as occurs in SJL, metabolic dysfunction can follow.

Our finding that higher household socioeconomic status (SES) significantly predicted baseline SJL seems logical, but it highlights an important pattern. Higher SES households often have greater access to electronic devices, which aligns with a systematic review and meta-analysis showing a positive association between SJL and screen media use [17]. The blue light emitted from these screens suppresses melatonin, the sleep-promoting hormone, and evening exposure is a major contributor to circadian phase delays[18].

Our study’s finding of the strong association between SJL and having a child aged 5-11 years is concerning as research has shown that in children aged 8-10 years, SJL was significantly associated with adverse cholesterol and vascular health factors, even when sleep duration was not affected, highlighting the unique and independent risk posed by mistimed sleep by itself [3]. This suggests that the metabolic consequences of SJL can manifest very early in life, setting a trajectory that may be difficult to change. Research shows that delayed weekend sleep patterns are already present in preschool children, suggesting parental influence has a foundational impact on a child’s sleep-wake pattern[11]. This is critical because SJL in school-aged children is associated with higher adiposity and emotional dysregulation[3,6]. Furthermore, a child’s temperament is shaped during these early years. While one cohort study found no direct association between SJL and temperament in pre-schoolers, it did find that increased sleep duration was associated with increased negative affectivity, a dimension linked to internalizing problems[11]. This complex interplay between sleep timing, duration, and a child’s developing personality underscores the profound importance of healthy sleep habits from a very young age.

The tournament served as a natural experiment, demonstrating how a compelling social event can function as a powerful, acute disruptor of sleep-wake cycles. The data show that a substantial proportion of children experienced additional social jetlag of additional one-hour sleep time displacement due to match watching; 33% on weekdays and 17.8% on weekends had. This provides compelling real-world evidence that social and entertainment demands can directly override biological sleep-wake drives, forcing children to be active and asleep outside their optimal circadian window[2].

This acute disruption has immediate consequences. A meta-analysis on sleep restriction and deprivation across the lifespan found that sleep loss has a large, diminishing effect on positive mood states and a moderate increasing effect on negative mood states, with these effects being particularly strong in younger people. Furthermore, sleep loss impairs the adaptive management of emotions[19]. This provides a plausible mechanistic explanation for the established links between SJL and behavioural problems such as emotional symptoms, hyperactivity, and conduct problems in pre-schoolers[6], as well as irritable mood and anxiety in adolescents[2].

The dramatically higher odds of SJL during FIFIA season reported by international parents warrant further investigation. This could be related to time zone differences with the primary match locations in Qatar, leading to even later and more disruptive viewing times. It may also reflect differing cultural norms around sleep hygiene, family media use, and the perceived importance of a consistent bedtime. These findings highlight that the drivers of SJL are not uniform and that interventions may need to be culturally tailored.

Children who had FIFA-related weekend sleep time delays were four times more likely to also have weekday delays. Our results clearly illustrate that sleep disruption is rarely confined to one part of the week; it creates a vicious cycle of compensation and instability. This ripple effect suggests that disrupting the structured schedule of a school night can destabilize the entire sleep-wake cycle, leading to the kind of high intraindividual variability that systematic reviews in adults have consistently associated with adverse health outcomes[13]. This irregularity is a hallmark of circadian disruption. The FIFA tournament, by driving later nights on weekdays, intensified this damaging cycle, making the re-synchronization of the circadian clock even more challenging.

While our study focuses on children, the implications of chronic SJL extend across the lifespan. The findings from adult populations provide a chilling preview of the potential long-term trajectory. In adults, variability in sleep habits between workdays and free days is common, with high percentages experiencing long-term sleep deprivation and chronic social jetlag, which are in turn associated with trouble sleeping and daytime sleepiness[20]. Longitudinal studies in adults have found associations between late chronotype, social jetlag, and worse mental and physical health outcomes, including mortality[21]. Evening types, who are most prone to SJL, appear to be cognitively more vulnerable and have a higher representation of mood disorders and attention deficit hyperactivity disorder (ADHD)[4]. The repeated need for temporal readjustment over the course of many years, a phenomenon our study shows can begin in early childhood, may negatively affect health in a broad sense[21]. Therefore, the high levels of SJL observed in our pediatric sample are not just a current concern, but a potential predictor of future chronic disease burden.

### Limitations and Strengths

The most significant limitation is its cross-sectional design, which captures data at a single point in time. This inherently prevents the establishment of causal relationships. For instance, while we can assert that FIFA-related sleep delays are associated with perceived sleep pattern changes, we cannot definitively prove that the tournament caused the change, as unmeasured confounding variables may be involved.

Secondly, the reliance on parental-reported data introduces the potential for several biases. Recall bias may affect the accuracy of habitual sleep schedules, and social desirability bias may lead parents to under-report problematic sleep behaviours or over-report their adherence to healthy sleep practices. Furthermore, parental perception may not always align with objective reality; a child’s internal state of sleepiness or circadian phase cannot be fully captured through mere observations. Additionally, part of our sample admitted that their children do not watch football matches basically which limits the association of their sleep habits and its disturbances with the FIFA season.

Lastly, concerns regarding generalizability must be acknowledged. Our sample predominantly consisted of highly educated parents residing in Saudi Arabia. The experiences and socioeconomic dynamics of this group may not be representative of more diverse populations, or those in different cultural contexts with varying attitudes toward sleep and media use.

Despite these limitations, the study possesses considerable strengths. Its large sample size provides robust statistical power and enhances the stability of the estimates derived from our multivariate models. The validity of investigating a real-world event (FIFA World Cup) is a significant advantage, as it captures sleep disruption within the context of genuine life. The application of multivariable logistic regression analyses strengthens the findings by controlling for a range of potential confounding sociodemographic factors thereby providing more reliable results. Lastly, the study’s focus on the intersection of a major social event and pediatric sleep fills a gap in the literature, providing unique insights into how acute social pressures can exacerbate a chronic public health issue.

## Conclusion

In conclusion, this study demonstrates that children have high baseline social jetlag. The FIFA World Cup, a major social event, additionally significantly disrupted their sleep schedules, acting as a potent catalyst for social jetlag, on top of already high baseline rates of chronic circadian misalignment. Children aged 5-11 years were clearly perceived highly vulnerable for social jetlag by their parents and should be targets for public health efforts.

Firstly, public health initiatives should provide proactive, practical guidance to families on managing sleep during large-scale events, emphasizing the importance of protecting morning light exposure and wake times, and strategically limiting late-night screen exposure for live matches in favor of recorded viewings when possible. More importantly, these findings add to the growing body of evidence calling for systemic changes that align social schedules with human biology. The most effective intervention would be to adjust school start times for children and adolescents to match their natural circadian rhythms. Such a policy would directly address the biological phase delay of puberty and help reduce the chronic SJL that undermines both the health and academic potential of an entire generation. Mitigating the long-term public health burden of SJL requires an intensive effort that addresses both the acute triggers and the foundational mismatch between our biological clocks and the demands of our social world.

## Data Availability

All data produced in the present work are contained in the manuscript.

## Disclosure

The authors declare no conflicts of interest.

## Acknowledgement

The authors thank all participants in this survey, including parents, care givers and data collectors. While preparing this manuscript, the authors used ChatGPT-5 to refine the language and grammar, then the authors reviewed and edited the content as needed and they take full responsibility for the content of the submitted manuscript. The authors are grateful to the Deanship of Scientific Research, King Saud University, for funding through the Vice Deanship of Scientific Research Chairs. We also thanks Hodhodata.com for their support in statistical analysis.

## Appendix

**Table 3:**
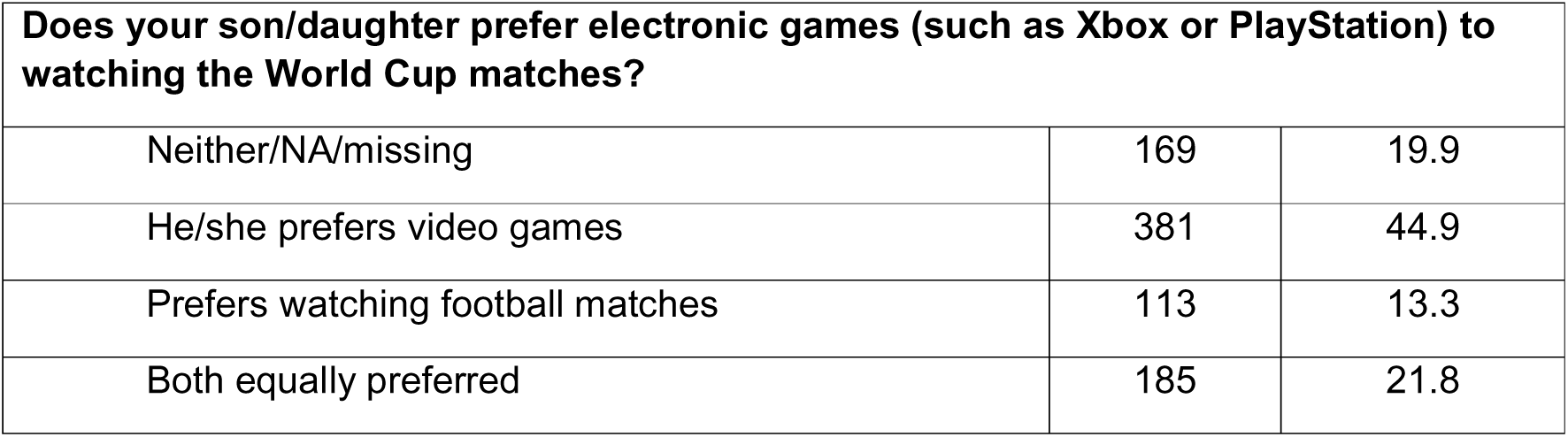
Parental perception of children behaviour of watching FIFA matches.

## Notes

### Competing Interest Statement

The authors have declared no competing interest.

### Funding Statement

This study did not receive any funding.

